# A Very Flat Peak: Why Standard SEIR Models Miss the Plateau of COVID-19 Infections and How it Can be Corrected

**DOI:** 10.1101/2020.04.07.20055772

**Authors:** Santosh Ansumali, Meher K. Prakash

**Affiliations:** Engineering Mechanics Unit, Jawharlal Nehru Center for Advanced Scientific Research, Jakkur, Bangalore 560064; Sankhya Sutra Labs Pvt Ltd, Manyata Embassy Business Park, Bangalore 560045; Theoretical Sciences Unit, Jawharlal Nehru Center for Advanced Scientific Research, Jakkur, Bangalore 560064; VNIR Biotechnologies Pvt Ltd, Bangalore Bioinnovation Center, Helix Biotech Park, Electronic City Phase I, Bangalore 560100

**Keywords:** COVID-19, Pandemic, Peak, Plateau, Asymptomatics

## Abstract

Innumerable variants of the susceptible-exposed-infected-recovered (SEIR) model predicted the course of COVID-19 infections for different countries, along with the ‘peaks’ and the subsequent decline of infections. One thing these models could not have predicted prospectively in January or did not adapt to in the following months is that the peak is rather a ‘plateau’ for many countries. For example, USA and UK have been persisting at the same high peak of approximately 30,000 and 5,000 daily new infections respectively, for more than a month. Other countries had shorter plateaus of about 3 weeks (6,400 cases in Spain). We establish that this plateau is not an artifact, and the “persistence number” describing the decline needs an equally important attention as the “reproduction number”. The solution lies in including the specific epidemiological role of asymptomatics and pre-symptomatics in COVID-19 transmission, different from SARS and influenza. We identify the minimal changes that can be made to any SEIR model to capture this plateau while studying seasonal effects, mitigation strategies, or the second wave of infections etc.

## I. INTRODUCTION

The World Health Organization had declared COVID-19 as a pandemic, the first one in the 21^st^ century [1]. The infections caused by the novel coronavirus (COVID-19) continue to increase beyond 4 millions and there are still many lacunae in the understanding of how it spreads and who are susceptible to the infection [2,3]. There are no known scalable treatment or vaccination options. SARS spread in around 29 countries, infecting around 8,096 individuals globally [4], and was contained with quarantine strategies [5]. The two natural scientific impulses in the face of the pandemic are to draw parallels to the containment measures and modeling efforts guiding them, that were successful in controlling SARS 2003 and MERS in 2015.

In response to COVID-19, Governments across the world had weighed the options of non-pharmaceutical interventions such as reduced social contact [6] and strict lockdowns [7-9]. The policy decisions had been motivated by epidemiological models that typically fall in the framework of Susceptible-Infected-Recovered (SIR) or Susceptible-Exposed-Infected-Recovered (SEIR) model or their variants [10,11], which for the sake of simplicity will be referred to as SEIR models for in this work. According to the SEIR models, in an unchecked spread [12] and using the early estimates for the basic reproduction number (R_0_=2.2) of COVID-19 [13] the spread of infections slows down when about 55% of the susceptible individuals have been infected. The hospital resources, health care personnel, and economies in many developed countries were overwhelmed when as little as 0.2-2% of the population was infected. Further, initially individuals over 65 years of age and comorbidities were considered highly susceptible. However, although at a much lower rate, healthy individuals in their 20s or 30s succumbed to COVID-19. Thus SEIR models focusing either on the entire population or only a specific demographic are both not practically relevant for COVID-19.

Because of the economic damage and the social inconvenience caused by the containment measures, public and policy makers alike were interested in crossing the ‘peak’ of daily infections [14]. Crossing was meant to send positive signals of pandemic containment to the people as well as to the economy, suggesting that time to lower the guard is not too far. However, the peak that was seen in many countries was a plateau – flat and lasting for many weeks – without showing a downward trend or a decline that is extremely slow compared to the predictions of the standard SEIR models. The first wave of the pandemic may finish soon, and even now there are no known therapeutic interventions. At least it is an opportunity to refine the modeling efforts that will continue for understanding and managing the second wave of COVID-19 infections or a chronic lower level of community infections have to understand the reason behind this serious oversight and adapt to it. We address this plateau in the minimal way using the known epidemiological information about the role of asymptomatic individuals, which is different from other well analyzed infectious diseases – influenza, SARS.

## II. RESULTS

### A. Many countries reach a plateau due to containment

**Fig. 1** shows the number of daily new infections from USA and UK. As it can be seen the new infections stop increasing, but persist almost at the same level for almost a month. The number of daily new infections is stochastic and one has to look for a moving average to be sure that the upwards, sideways or downwards trends are not incorrectly interpreted. However, even after such moving average, the trend shown by the daily new cases in **Fig. 1** is very different from the classic picture of the peak described by the SEIR models [12,14] where the number of cases decline soon after crossing the peak, almost in a quasi-symmetric reflection of the phase when the cases increased. This flat peak or a plateau in the number of daily new infections reflects as a persistent linear increase in the number of cumulative infections, as could be seen (**Supplementary Fig. 1**) for South Korea, Switzerland, Spain, Germany [15] as well as some of the individual provinces or cities – New York city, Bayern (Germany), Veneto (Italy), Catalonia (Spain). The slope of this linear growth phase region indicates the average number of daily new cases. The linear phase lasted much shorter in many countries, for only about 21 days. When studying and interpreting the data from a heterogenous society such as a large country, where each state could have different containment policies, it is important to decouple the data from the regions as it allows a more meaningful and early detection of the trends. For example, one region could be still be in an exponential growth phase and another on the plateau with a linear growth rate, the overall numbers can be confusing and hard to interpret. **Fig. 2** shows the data from one of the epicenters in Italy, the Lombardy region [16]. By decoupling it from the pandemic evolution in Italy, we could identify the linear growth early on, within a few days after the lockdown was imposed on March 9, 2020.

**Fig. 1.**
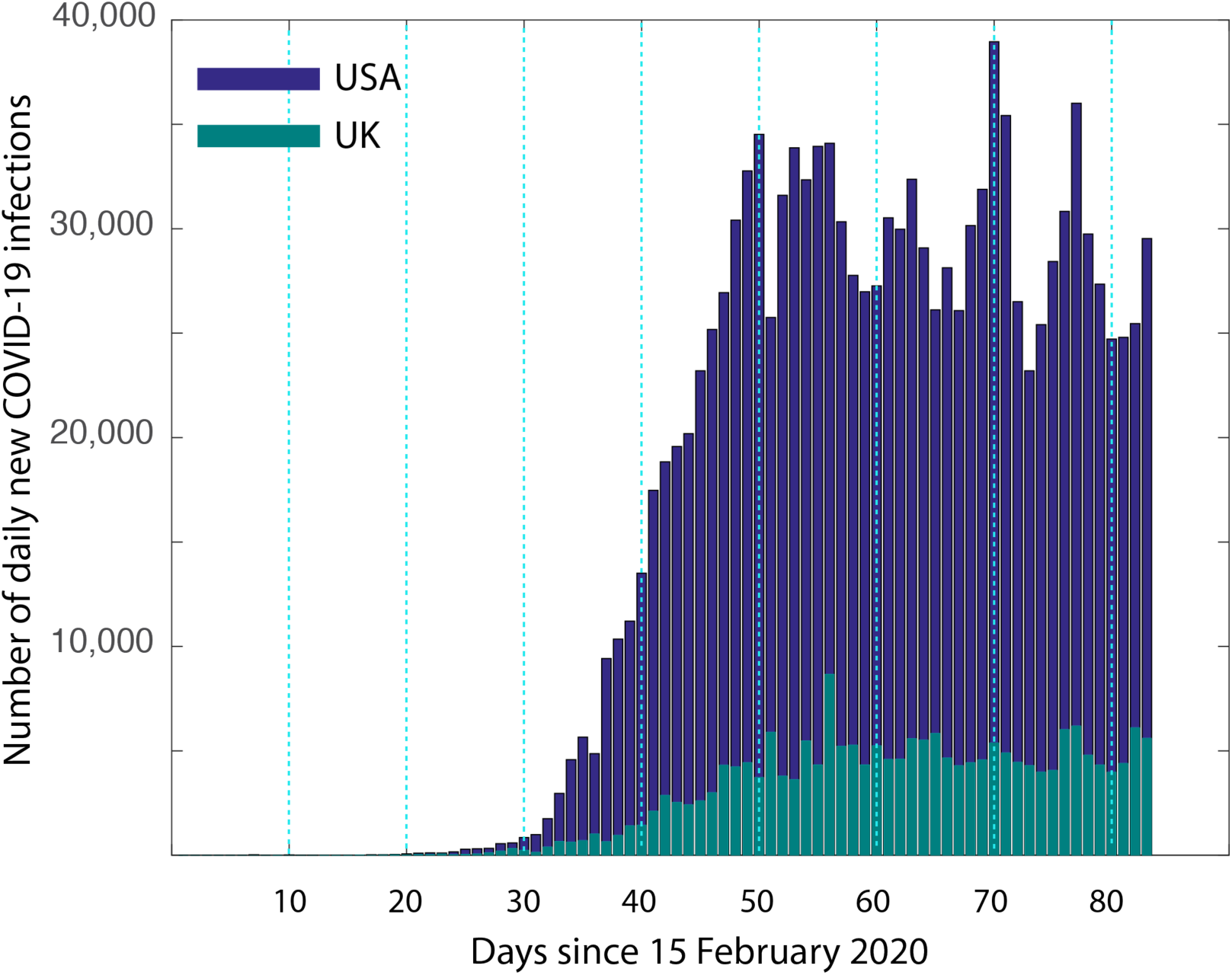
Plateau in the number of daily infections. The number of daily infections (February 15-May 7) from USA and UK are shown. It is clear that neither of the two shows a classic quasi-symmetric peak that is predicted by the standard SIER models [12]. So it is rather appropriate to call it a “flat peak” or a “plateau”. Although the predictions of the beginning of the peak may have been correct, this partial agreement which does not suggest the possibility of a plateau can be mislead one into believing that the cases after the peak will decline.

**Fig. 2.**
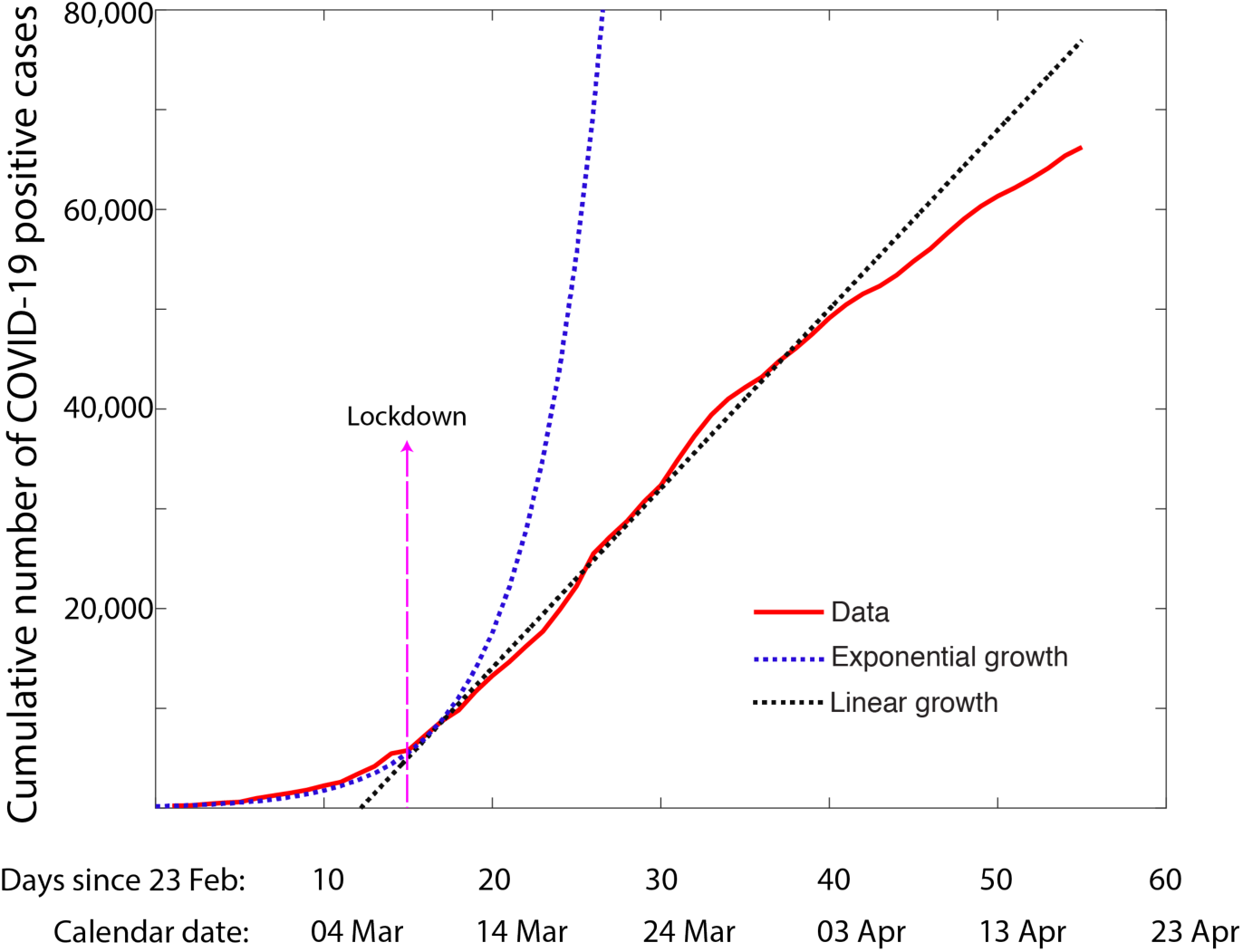
Exponential to linear transition in Lombardy province of Italy. The cumulative infections which show an exponential transition continue with an average of 1974 new COVID-19 positive cases. The date on which the lockdown was implemented is shown. Interestingly, the transition to linearity happens within around 10 days after the lockdown.

### B. Interpreting the Plateau

#### Relief from exponential growth

The significance of a deviation from the exponential to non-exponential growth rate is that there are no new infections are caused by an individual who is understood be infected (d*I*/dt ~ *I*, where *I* is the number of cumulative infections results in an exponential growth). A deviation from an exponential growth phase whether the immediate result is a peak or a plateau in daily new cases is a sign of success of the containment measures [17], and a relief to the population. In most places, this deviation from the exponential growth was achieved within a week to 10 days after the implementation of the containment strategies, possibly coinciding with the incubation time of COVID-19, an epidemiological term to describe the time between the infection and the onset of symptoms [18].

#### Pause and not a stop

In this plateau of the number of infections, many countries showed very high number of daily new cases for many weeks. The daily new infections in Switzerland (959), Germany (≈5260), Spain (≈6938) despite the best containment measures (**Table 1**) they could adopt. The continued new infections cannot be because of the incubation period. The incubation period has been estimated to be with a mean of 6.4 days [18], with about 95% completion of incubation by 15 days. They cannot be because of the excessive testing either. Other than Germany or South Korea, the average in most countries was below testing 3 individuals for every one detected positive (**Table 1**). Even many developed countries did not test the family members living in the same household as an infected individual, unless they self-reported symptoms. Thus the plateau where there is a steady level of new infections signifies a phase where the system is held in a pause with the potential to spring back to exponential phase.

**Table 1.** Data showing the average number of daily new infections (slope in the linear regime) along with other relevant information for each country or province. ^*^Estimated from the German national testing as of 1 April 2020. ^§^The infection data from South Korea shows two different linear regimes. The first one which immediately follows the end of the exponential regime is considered.

|  | Date of lockdown | Cumulative infections (on the date of lockdown) | Tests/Each detected infection | Population (Millions) | Average daily new infections (slope) |
| --- | --- | --- | --- | --- | --- |
| Lombardy | 9 March | 4,224 | 2.6 | 10 | **1797** |
| Veneto | 9 March | 715 | 11.6 | 4.9 | **420** |
| Bayern | 21 March | 3,650 | 14.1* | 13 | **1372** |
| Catalonia | 14 March | 715 | 1.92 | 7.6 | **1432** |
| New York City | 22 March | 9,654 | 1.70 | 8.4 | **4855** |
| Spain | 14 March | 6,391 | 1.92 | 46.7 | **6938** |
| Germany | 21 March | 22,213 | 14.1 | 82.8 | **5261** |
| South Korea | 29 February | 3,150 | 43.3 | 51 | **616<sup>s</sup>** |
| Switzerland | 17 March | 3,028 | 7.6 | 8 | **959** |

#### Daily cases depending on the lockdown date

The daily new infected cases for the provinces and countries we analyzed in **Figs. 2, 3** are given in **Table 1**. The dependence of the daily new cases on various factors such as the extent of testing, the population of the region, etc was studied in **Fig. 3**. A correlation was observed with the number of cumulative infections in the region, and the number of daily infections at the time when the containment measures were taken. Which suggests that implementing the lockdown a few days later in the exponential growth phase, has two effects – the number of cases increase over these days, but more importantly for the plateau to follow the number of daily infections for a many weeks to months will scale with the number of infections on this later date. Since the median recovery time of individuals infected by COVID-19 is around 3 weeks, the new infections will continue to burden the healthcare system. This ‘exponential mistake’ will have a continued effect with high number of daily infections, presenting a strange situation of having chronic and acute severity simultaneously for a long time.

**Fig. 3.**
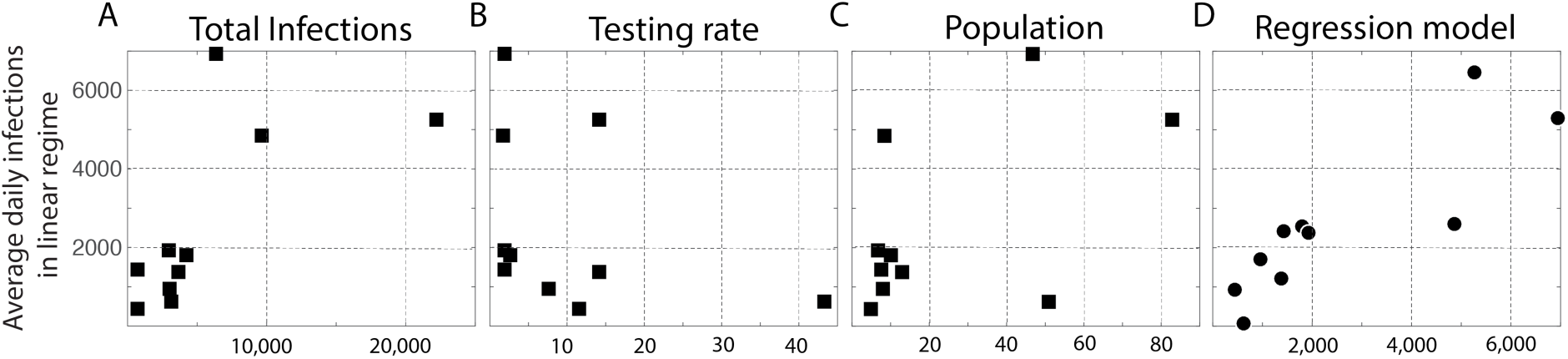
Factors correlating with the constant average daily infections. The slope from the linear regime was compared with several factors to see if any factor could potentially help interpret the linear regime. (A) The cumulative number of infections at the time of the lockdown, (B) Testing rate, which is the number of tests performed for every detected positive, (C) Population of the region (millions) were are all compared with the average daily infections (y-axis). The number of infections at the time of the lockdown seemed to have the best correlation among the three. As much as the linear regime suggests the end of the exponential growth phase, a correlation of the daily cases with the average number of infections at the time of transition seems to suggest that the growth is maintained in a “pause”, frozen at the state where the quarantines are implemented. D. A linear regression model for combining the factors in A, B, C (=0.0127 * Number of Infections at lockdown −133 * Tests per positive + 71.92*Population in millions+2110) was used to predict the number of daily infections, and clearly the correlation with this combination of factors improves.

### C. Model elements to capture the unique character of COVID-19

After noticing that the plateau is not an artifact of excessive testing or the incubation, depends significantly on the point when the lockdown was implemented and hence cannot be ignored, one wonders why this has not been captured in the many variants of SEIR models [12, 14]? There are already several flavors of SEIR models. Some of the very detailed models capture very slow decrease in the cases, but the mechanics of slow decay remains buried in the extremely detailed models, mathematics and has not been discussed [19]. Our objective is not to create one more, but to identify the minimal elements of COVID-19 specific epidemiology that can capture the observed plateau and hence can be integrated into any SEIR model.

Reviewing the understandings into the epidemiology of COVID-19, there are at least three unique characteristics: The individuals who will eventually become symptomatic have a 44% chance of spreading the infections before becoming symptomatic [20]. Further, there around 86% of the infected population is expected to remain asymptomatic [21], and yet contribute to the spread of infections [21]. Interestingly, the infectiousness profile of SARS-CoV-2 is very different from that of that of SARS-CoV-1 which caused SARS [22] and of influenza [23]. Unlike SARS-CoV-2, these two respiratory diseases have a peak of infectiousness after the onset of symptoms (**Fig. 4**). Thus a spread of infections mainly through the symptomatic individuals is justified for these diseases, but not for COVID-19, especially in an imperfect lockdown where the unsuspecting asymptomatic and pre-symptomatic spread the new infections. SEIR variants including the possibility of the spread of infections by asymptomatic individuals exist [24], however to the best of our knowledge have not gained relevance in COVID-19 epidemiology.

**Fig. 4.**
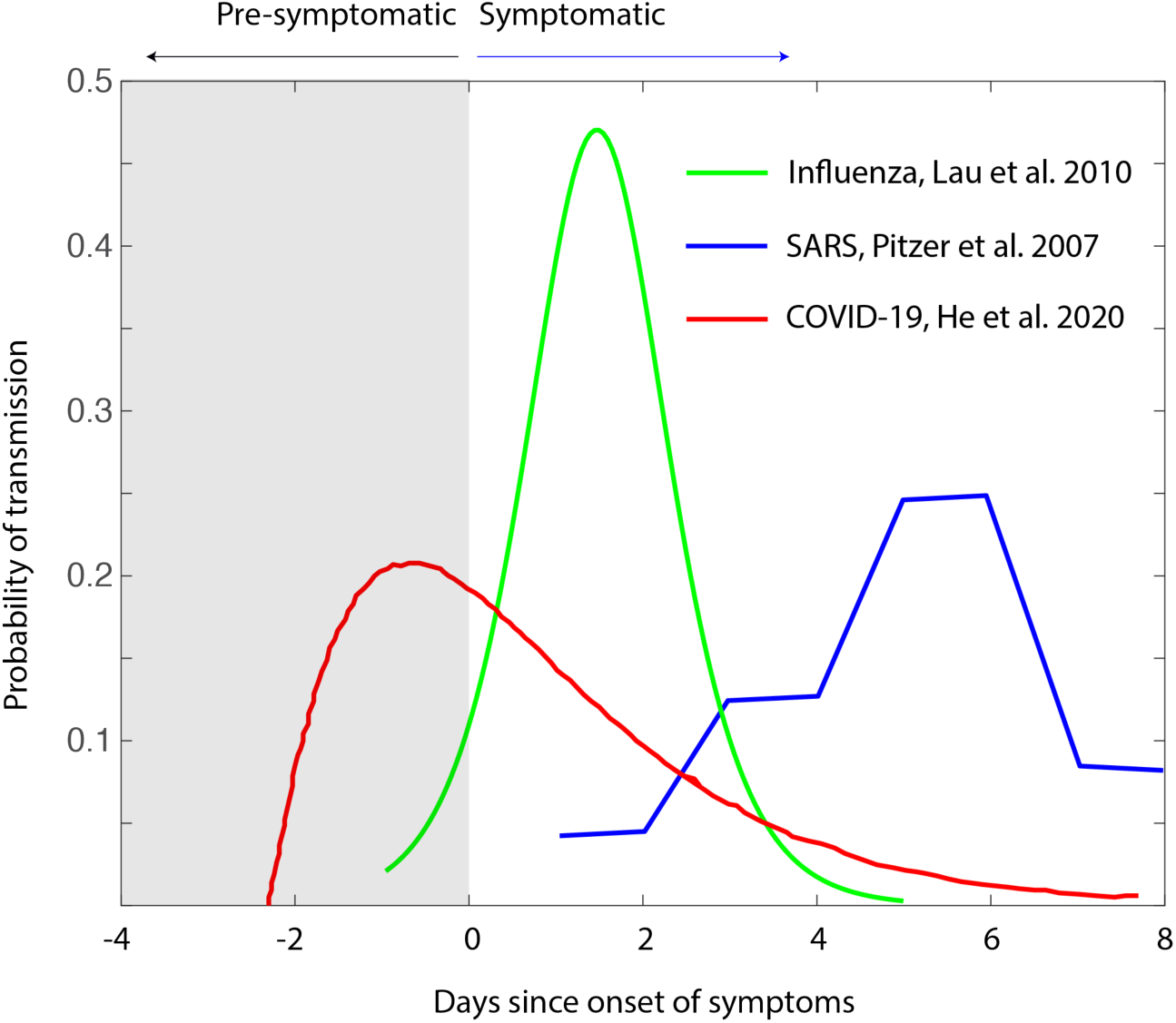
Temporal infectiousness profiles. The differences in the infectiousness profiles of influenza [22], SARS [21], and COVID-19 [19] are shown. The gray box indicates the period before the onset of symptoms. The graphic highlights the clear differences of spread of infections via pre-symptomatic individuals, needed to be noted in modeling COVID-19 relative to the models used for influenza, and even for its close homolog SARS-CoV-1 which caused the SARS of 2003.

Our minimal model for the evolution of the number of infected (*I*), asymptomatic (*A*) which also includes the pre-symptomatic in this work, is as follows:

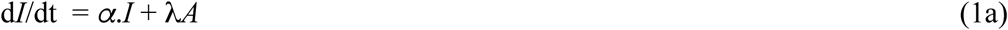

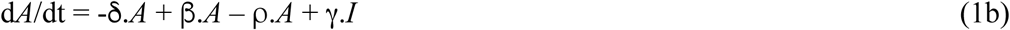

*α* is the transmission of infection by infected individuals, which implicitly includes the susceptible population that does not change significantly. λ is the infection rate via asymptomatic individuals, δ reflects the natural rate of reduction in the numbers of asymptomatic patients post-incubation period, β is the rate at which asymptotic individuals transmit to other individuals, *ρ* is the rate at which by performing tests one reduces the *A* by moving them to a quarantine and γ is the rate at which infected individuals, in an imperfect quarantine can spread new infections resulting in asymptomatics. A median hospitalization time of 3 weeks from the data does support this assumption, as in the exponential phase the increase in the number of infections in these 3 weeks is much higher. The decrease of infections due to recovery that is slow and increase of infections due to non-human sources such as aerosols or contact surfaces have not been considered.

When the lockdown is implemented, and no exponential growth could be seen, and at least that the infected individuals are perfectly quarantined, α=0 and γ=0 respectively can be assumed. In **Supplementary Fig. 3**, we illustrate a simulation of these differential equations to show the transition to a flat plateau for the chosen parameters. The model can capture all the features that **Fig. 3** shows. But given the simplicity of the equations, one can also perform a fixed-point analysis. Considering **Eq. 1b**, the ratio β/(ρ+δ) will determine whether *A*, which continues to cause new infections in our assumption decreases with time, remains stable or increases with time going back to another kind of exponential dependence. The decrease in new infections can be more rapid if the testing ρ is high or the unwanted transmission β is low. This ratio is similar to the R_0_ that has been studied [24]. However when applied in the context of a lockdown where the cases are not declining, it gains a better significance when referred to as the persistence number. While it may seem a matter of semantics, referring to it as the persistence shifts the emphasis shifts from describing the capability of virus to spread to how fast the new cases decrease, in a way that can be quantified and compared to other societies.

## CONCLUSIONS

In summary, the aim of this work is to address the ‘elephant in the room’ that is the plateau of infections that appears in the observations but has not been addressed either by the statistical models or by SEIR models. We show that using the known epidemiology of the COVID-19, the infection models need to account for pre-symptomatic and asymptomatic, infections, something that was not equally relevant while modeling seasonal influenza or even SARS of 2003. We also underscore a way of plateau declines with a newer terminology, such as the ‘persistence number’ which actually shifts the emphasis from the nature of the virus to spread to the ability to control.

## Data Availability

All data will be available on request

## Acknowledgements

S.A and M.K.P acknowledge the support of JNCASR for this research. M.K.P acknowledges the discussions with Nadia Ambrosi and Yousung Jung which were very helpful in understanding the ground realities in Europe and South Korea. S.A. and M.K.P acknowledge Prof. K. R. Sreenivas, Dr. Sunil Sherlekar for helpful discussions.

## SUPPLEMENTARY FIGURES

**Supplementary Fig. S1.**
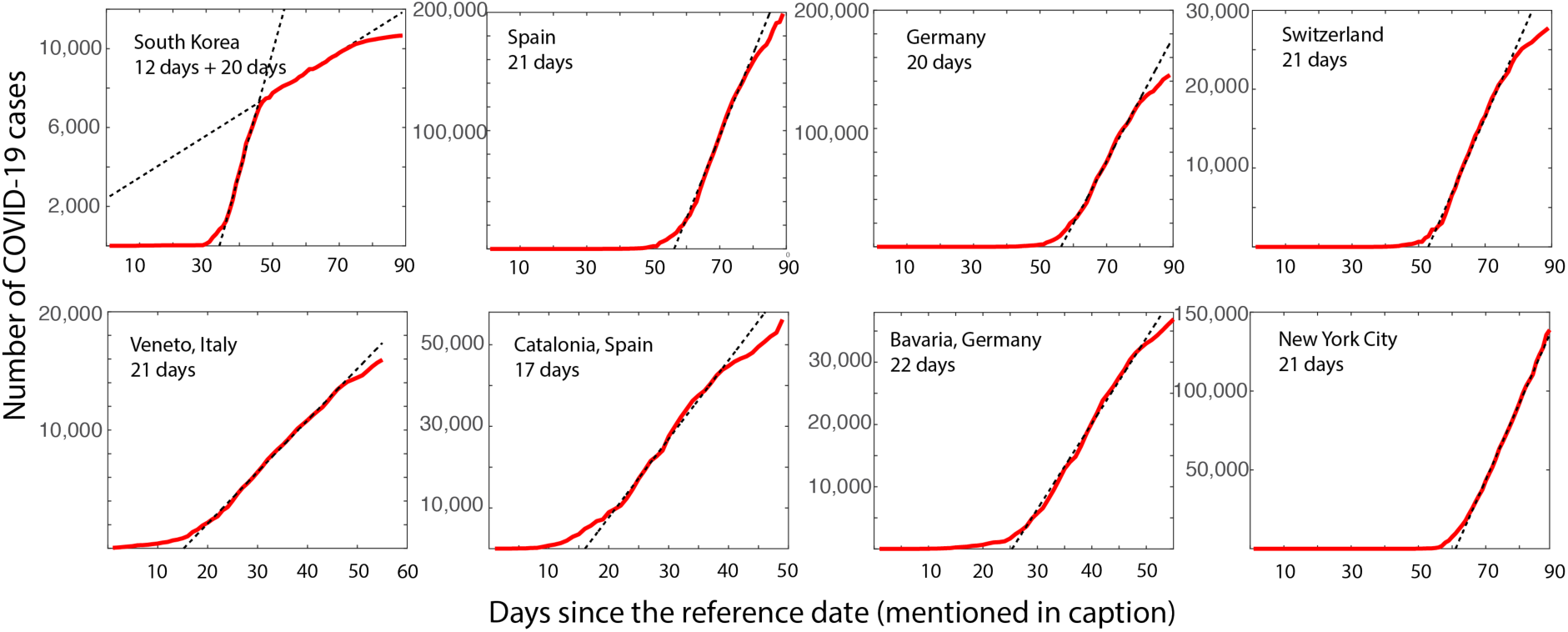
Transition from exponential to linear in several countries and regions. The plots show the infections in countries (top row) and provinces or cities (bottom row). As it can be seen the number of infections enters a linear regime for all these regions, and the corresponding slopes are given in **Table 1**. The data up to the 19^th^ of April was used in these analyses. The legends in the figures suggest that the number of days for which the growth continued in a linear regime. South Korea have two distinct linear regimes, and the duration in both are indicated. x-label for Veneto, Bavaria are days since 24 February 2020, Catalonia from 28 February 2020 and all other data is days since 22 January 2020.

**Supplementary Fig. S2.**
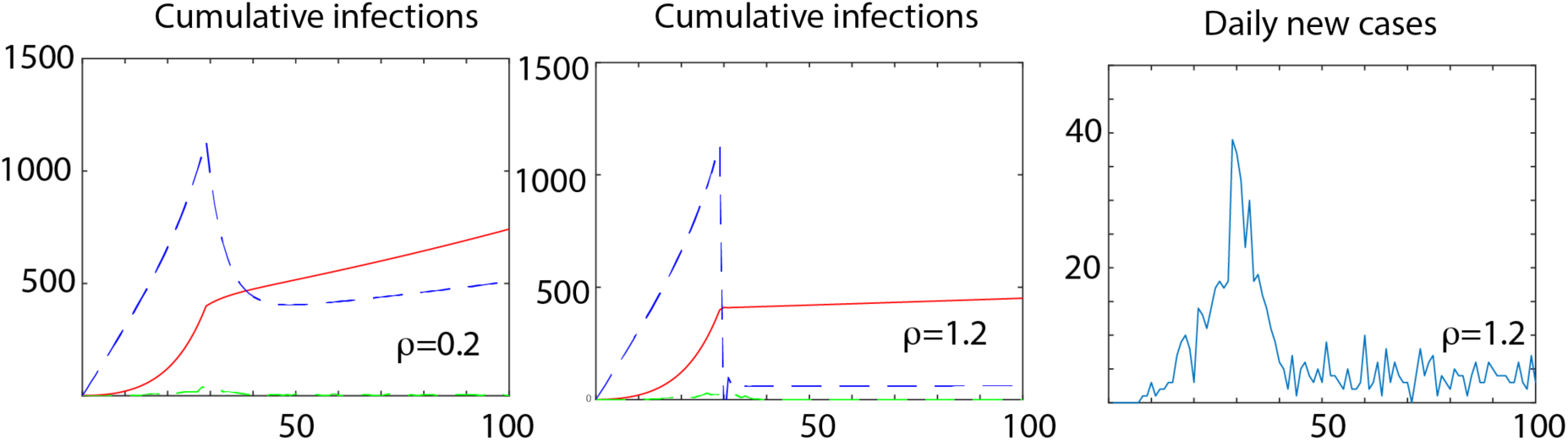
Model predictions of the combined Infected vs. Asymptomatic model. The rate equations in Eq. (1) were simulated to see if the observations can be captured. The simulation was performed with parameters such as α=0.1, and the transmission via asymptomatic is individuals is much lesser with λ=0.01. At this growth stage of the pandemic since the data from most countries show a recovery rate of around 10%, we performed the simulation with μ=0. The asymptomatic (blue), and recorded infections (red) with two different testing rates ρ=0.2 and 1.2 initiated after the lockdown period (30 on axis which represents the days) are shown in panels A and B. As expected a transition from an exponential to linearity is observed. Panel C shows the daily new infections when ρ=1.2. Since the Equation (3) is stochastic, we added an incubation period drawn randomly from a beta-distribution with a mean of 6.5 days. For this choice of parameters which were chosen to qualitatively emulate the peak observed in South Korea, a low level of daily new infections persist. However, by varying ρ, it was seen that these average number of these daily new cases decreases with the r and increases with the time of lockdown (data not shown).

